# Post-discharge mortality among children under 5 years admitted with suspected sepsis in Uganda: a prospective multi-site study

**DOI:** 10.1101/2023.01.12.23284164

**Authors:** Matthew O Wiens, Jeffrey N Bone, Elias Kumbakumba, Stephen Businge, Abner Tagoola, Sheila Oyella Sherine, Emmanuel Byaruhanga, Edward Ssemwanga, Celestine Barigye, Jesca Nsungwa, Charles Olaro, J Mark Ansermino, Niranjan Kissoon, Joel Singer, Charles P Larson, Pascal M Lavoie, Dustin Dunsmuir, Peter P Moschovis, Stefanie Novakowski, Clare Komugisha, Mellon Tayebwa, Douglas Mwesigwa, Cherri Zhang, Martina Knappett, Nicholas West, Vuong Nguyen, Nathan Kenya Mugisha, Jerome Kabakyenga

## Abstract

**Background:** Substantial mortality occurs after hospital discharge in children under 5 years old with suspected sepsis. A better understanding of its epidemiology is needed for effective interventions aimed at reducing child mortality in resource limited settings.

**Methods:** In this prospective observational cohort study, we recruited 0-60-month-old children admitted with suspected sepsis from the community to the paediatric wards of six Ugandan hospitals. The primary outcome was six-month post-discharge mortality among those discharged alive. We evaluated the interactive impact of age, time of death, and location of death on risk factors for mortality.

**Findings:** 6,545 children were enrolled, with 6,191 discharged alive. The median (interquartile range) time from discharge to death was 32 (10–92) days, with a six-month post-discharge mortality rate of 5·5%, constituting 51% of total mortality. Deaths occurred at home (45%), in-transit to care (18%), or in hospital (37%) during a subsequent readmission. Post-discharge death was strongly associated with weight-for-age z-scores < -3 (adjusted hazard ratio [aHR] 5·04; 95%CI: 3·97–6·37), referral for further care (aHR 9·08; 95%CI 6·68–12·34), and unplanned discharge (aHR 3·36; 95%CI 2·64–4·28). The hazard ratio of those with severe anaemia increased with time since discharge, while the hazard ratios of discharge vulnerabilities (unplanned, poor feeding) decreased with time. Children with severe anaemia (<7 g/dL) died 35 days (95%CI 19·4–51·9) later than those without anaemia. Age influenced the effect of several variables, including anthropometric indices (less impact with increasing age), anaemia (greater impact), and admission temperature (greater impact).

**Interpretation:** Paediatric post-discharge mortality following suspected sepsis is common, with diminishing, though persistent, risk over the 6 months after discharge. Efforts to improve post-discharge outcomes are critical to achieving Sustainable Development Goal 3.2 (ending preventable childhood deaths under 5 years of age).

**Funding:** Grand Challenges Canada (#TTS-1809-1939), Thrasher Research Fund (#13878), BC Children’s Hospital Foundation, and Mining4Life.

## Introduction

Though substantial improvements in child mortality have been achieved over the past several decades,^1^ mortality following hospital discharge has emerged as a key priority to further improve child survival, especially in low-income countries. The successful transition of care between facility and community is a critical component of comprehensive care for acute illness and should not be decoupled from hospital care, as is often the case.^2^ A more granular understanding of events occurring during the post-discharge period is urgently required to guide interventions and policy decisions regarding resource allocation.^3^

Robust epidemiological data for paediatric post-discharge mortality in low-income countries has been limited, though a few studies have been published recently.^2,4–6^ Mortality following discharge results from the complex convergence of a series of causal factors, elegantly outlined in the recent CHAIN study.^5^ This suggests that post-discharge deaths involve a heterogeneous set of circumstances and that efforts to improve survival following discharge must entail interventions across several domains.

Consistently reported features of those at risk of post-discharge mortality include malnutrition, illness severity (at admission and discharge) and socioeconomic factors such as poverty and maternal education.^2,7^ Significant epidemiological gaps remain, however, including how age interacts with known risk factors. Furthermore, while several studies have reported high levels of at-home deaths,^2^ risk factors for these deaths is poorly understood. Finally, while many deaths occur early during the post-discharge period, it is unclear how host factors at admission impact the timing of post-discharge deaths. These data are imperative in building prediction models and interventional programs that address the complexities of paediatric post-discharge mortality.

The aim of this study is to improve our understanding of the heterogeneous nature of post-discharge mortality, by examining its incidence by age, location of death, and time-of-death among children under-5 admitted to hospital with suspected sepsis in Uganda.

## Methods

### Study design

This study analysed post-discharge outcomes from a combined dataset comprising two prospective observational cohort studies of children with suspected sepsis, aged 0-6 months and 6-60 months at the time of admission to a hospital in Uganda. This study was approved by the Mbarara University of Science and Technology Research Ethics Committee (No. 15/10-16), the Uganda National Institute of Science and Technology (HS 2207), and the University of British Columbia / Children & Women’s Health Centre of British Columbia Research Ethics Board (H16-02679). Written informed consent was obtained from a parent or guardian prior to the enrolment of eligible study subjects. This manuscript adheres to the guidelines for STrengthening the Reporting of OBservational studies in Epidemiology (STROBE).^8^

### Setting

The study enrolled participants from six hospitals in Uganda: Mbarara Regional Referral Hospital (Southwestern Uganda), Holy Innocents Children’s Hospital (Southwestern Uganda), Masaka Regional Referral Hospital (Central Uganda), and Jinja Regional Referral Hospital (Eastern Uganda). Children less than 6 months were also enrolled at Villa Maria Hospital (Central Uganda) and Uganda Martyrs Hospital, Ibanda (Southwestern Uganda). In total, these facilities have catchments covering 30 districts with a total population of over 8 million individuals, including approximately 1·4 million under-5 children during the study period,^9^ which provides a reasonable representation of the national Ugandan population outside Kampala.

### Study populations

This study involves two cohorts: the first cohort enrolled children 6-60 months and the second enrolled children 0-6 months of age. Both studies were conducted by the same investigative team who used the same research staff to enrol and follow up all participants. Enrolment of each cohort was independently funded.

We enrolled children with suspected sepsis, defined as children who were admitted with a proven or suspected infection, as determined by the treating medical team. We have previously demonstrated that approximately 90% of children admitted to hospital with a proven or suspected infection meet the International Pediatric Sepsis Consensus Conference (IPSCC) definition for sepsis.^10^ Enrolment occurred July 2017 to July 2019 (6–60-month cohort) and January 2018 to March 2020 (0-6-month cohort), with follow-up occurring until 6 months after hospital discharge. Children who resided outside the hospital catchment or who were admitted for a short-term (<24 hour) observation period, trauma, or immediately after birth (i.e., without first being discharged home) were excluded.

### Data collection procedures

All data collection tools are available through the Smart Discharges study Dataverse.^11^ All data were collected at the point of care using encrypted study tablets and these data were then uploaded to a Research Electronic Data Capture (REDCap)^12,13^ database hosted at the BC Children’s Hospital Research Institute (Vancouver, Canada).

At admission, trained study nurses systematically collected data on clinical, social and demographic variables. Clinical data included anthropometry (to determine malnutrition status), vital signs, simple laboratory parameters (glucose, malaria rapid diagnostic test [RDT], HIV RDT, haematocrit, lactate), clinical signs and symptoms, co-morbidities, and healthcare history, including previous hospital admissions. Social and demographic variables included maternal and household details.

Following discharge, field officers contacted caregivers at 2 and 4 months by phone, and in-person at 6 months, to determine vital status, any occurrence of post-discharge health-seeking, and readmission details. Verbal autopsies were conducted for children who had died following discharge (**Supplementary Table S1**).

### Statistical analysis

Target samples of 2,700 children 0-6 months and 3,500 children 6-60 months were based on the sample size required to develop two separate, age-specific, post-discharge mortality prediction models for cohorts with an expected outcome rate of 7·5% and 5%, respectively. These models will be reported in a separate publication.

For this analysis, data from both cohorts were combined and analysed as a single dataset. We used periods of overlapping enrolment (72% of total enrolment months) between the two cohorts to determine site-specific proportions of children who were 0-6 and 6-60 months of age (**Supplementary Tables S2, S3**). These proportions were used to weight the cohorts for the calculation of overall mortality rate.

Age-stratified Kaplan-Meier survival curves were used to estimate the cumulative hazard for overall and post-discharge mortality according to 4 pre-defined age strata (0 to <2 months, 2 to 6 months, >6 to 24 months, >24 to 60 months). Multivariable cox proportional hazard models, adjusted for age, sex, and site of enrolment, were fitted to estimate the impact of clinical, social, and demographic variables on post-discharge mortality. Logistic regression with interaction terms between age and other predictor variables of interest was used to assess possible age-specific effects (interactions) with mortality. No adjustment for multiple comparisons was made in these exploratory analyses.

To examine the changing risk over time among individual variables we divided the 6-month post-discharge period into tertiles, dividing post-discharge deaths evenly between these periods. Highly significant predictors (p-value <0·01) from the main cox regression model were selected and three hazard ratios for post-discharge death were derived, one for each tertile period. A heatmap was produced to visualise the magnitude of change in hazard ratios across the three periods, using the first period as the reference. The same set of predictors were used to determine their effects on time to post-discharge death by fitting a linear regression with a robust standard error with the number of days to death since discharge as the continuous outcome variable.

To determine the impact of predictors on location of death, a multinomial regression was fitted with location of death as the categorical outcome variable. The categories were: dying in hospital at readmission (the reference category), at home, or in transit to seeking care. A relative risk ratio was computed along with the corresponding 95% confidence interval (CI) and p-value. Only data from children who died post-discharge were used to identify factors associated with timing and location of death.

Missing values were very low and were imputed using k-nearest neighbours in all analyses. Analyses were conducted in Stata version 15.0/MP (StataCorp, College Station, TX), R version 4.1.3 (R Foundation for Statistical Computing, Vienna, Austria), and RStudio version 2022.2.3 (RStudio, Boston, MA).

## Results

Between July/2017 and March/2020, 16,991 consecutively admitted children were screened and 6,545 were enrolled: 2,707 into the 0-6-month cohort and 3,838 into the 6-60-month cohort (**Figure 1**).

**Figure 1:**
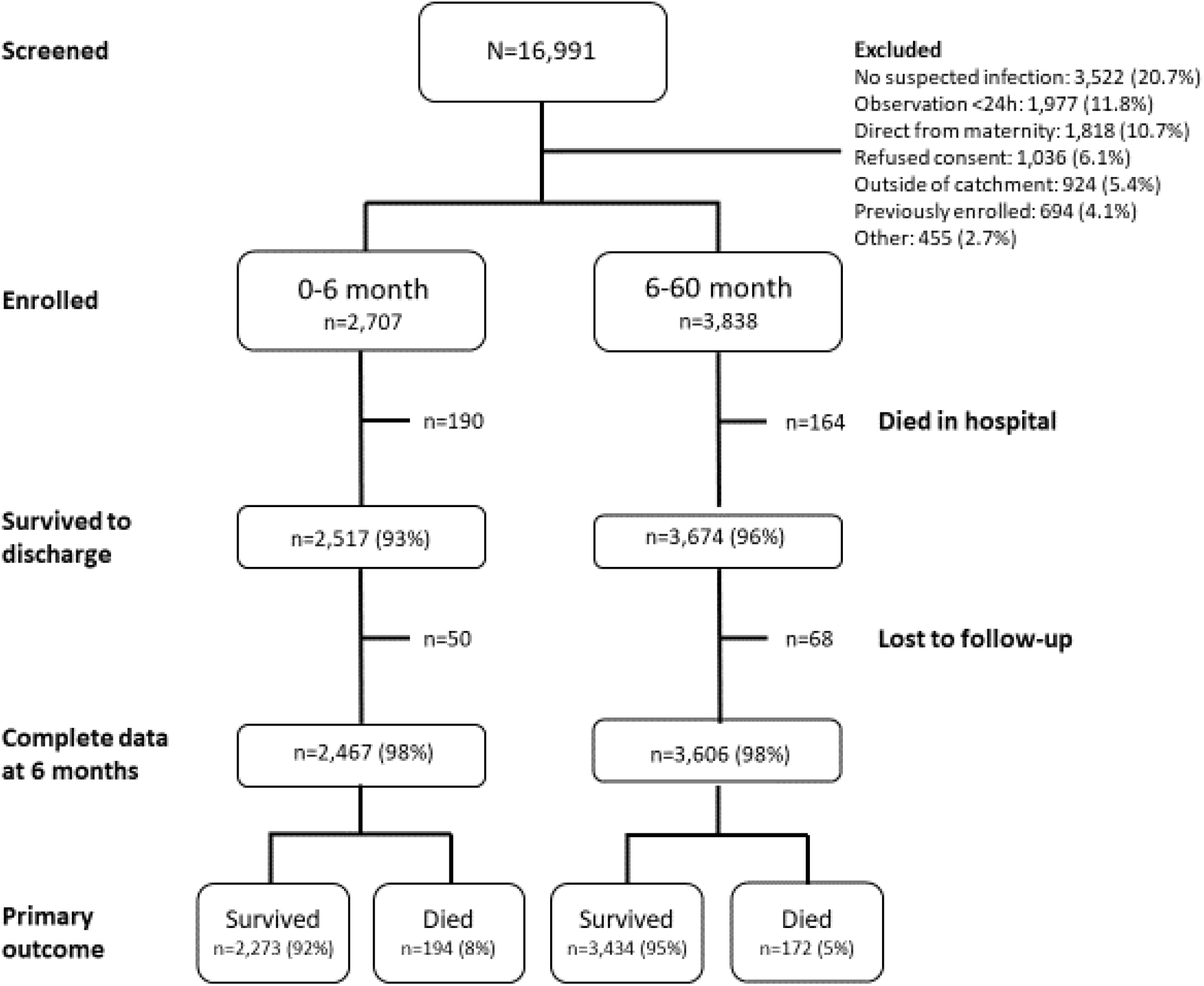
Flowchart of study screening, enrolment and follow-up for the two cohorts (0-6 month and 6-60 month)

### Mortality rates and associated diagnoses

Overall, the weighted 6-month mortality rate (in-hospital and post-discharge) was 10·9%. The overall median (interquartile range [IQR]) duration between admission and death was 9 (2–45) days.

6,191 children were discharged from hospital alive, of whom 118 (1·9%) were lost to follow-up at six months. Among these 6,191 children, the weighted six-month post-discharge mortality was 5·5%, which constituted 51% of total mortality. Of 361 verbal autopsies completed for post-discharge deaths, pneumonia (26·6%), malaria (8·6%), diarrhoea (8.0%), and meningitis (7·5%) were the most common specific diagnoses assigned (**Supplementary Table S1**).

Suspected sepsis deaths were also common (22·4%) and represented cases where a generalised infection was suspected.

### Risk predictors for post-discharge mortality

Malnutrition was commonly identified at admission and conferred significant risk for post-discharge mortality, with all anthropometric indices associated with significantly increased risk (**Table 1a**). A weight-for-age z-score less than -3 had an adjusted hazard ratio (aHR) of 5·04 (95%CI 3·97–6·37) for post-discharge mortality, with 13·5% of children in this stratum. Most clinical signs and symptoms pertaining to more severe illness on admission conferred increased risk for post-discharge mortality, although higher admission axillary temperatures were associated with a lower risk (**Table 1b**). Children with a positive rapid HIV antibody test were also at higher risk of post-discharge mortality (aHR 1·97; 95%CI 1·25–3·11), though children with a positive malaria RDT had a lower risk (aHR 0·73; 95%CI 0·55–0·98). Other factors associated with an increased risk included severe anaemia (aHR 2·81; 95%CI 2·08–3·81), elevated lactate (aHR 1·09; 95%CI 1·04–1·13, per 1 mmol/L increase), and hypoglycaemia (aHR 1·85; 95%CI 1·14–3·02). Several factors reflecting social vulnerabilities were also associated with increased risk of post-discharge mortality, including lower maternal education, increased distance from hospital (both time and actual distance), maternal HIV, and not using boiled, disinfected, or filtered water (**Table 1c**).

**Table 1.** Clinical risk factors for 6-month post-discharge mortality (N=6187): (a) demographic variables; (b) clinical assessment at admission; (c) maternal and social characteristics; and (d) discharge characteristics.

|  | n (%) / mean (SD) | aHR (95%CI) |
| --- | --- | --- |
| **a) Demographic variables** |  |  |
| **Male Sex** | 3446 (55.70) | 0.95 (0.77–1.17) |
| **Age (years)** | 1.11 (1.14) | 0.90 (0.82–1.00) |
| **Admission Anthropometry** |  |  |
| **MUAC** | 128.45 (20.56) | 0.96 (0.95–0.97) |
| <110/<115 | 1218 (19.69) | 5.54 (4.12–7.45) |
| 110-120/115-125 | 1135 (18.34) | 2.31 (1.66–3.22) |
| >120/>125 | 3834 (61.97) | ref |
| **Weight for age Z-scores** | -1.21 (1.79) | 0.72 (0.69–0.76) |
| <-3 | 838 (13.54) | 5.05 (3.97–6.43) |
| -3 to -2 | 855 (13.82) | 2.36 (1.77–3.16) |
| >-2 | 4494 (72.64) | ref |
| **Length for age Z-scores** | -0.79 (2.05) | 0.75 (0.72–0.79) |
| <-3 | 748 (12.09) | 3.99 (3.12–5.08) |
| -3 to -2 | 792 (12.80) | 2.07 (1.54–2.77) |
| >-2 | 4647 (75.11) | ref |
| **BMI Z-scores** | -1.04 (2.09) | 0.83 (0.80–0.87) |
| <-3 | 956 (15.45) | 2.78 (2.17–3.55) |
| -3 to -2 | 802 (12.96) | 2.05 (1.55–2.72) |
| >-2 | 4429 (71.59) | ref |
| **Weight for length Z-scores** | -1.04 (2.15) | 0.88 (0.84–0.92) |
| <-3 | 970 (15.68) | 2.15 (1.68–2.76) |
| -3 to -2 | 789 (12.75) | 1.57 (1.16–2.12) |
| >-2 | 4428 (71.57) | ref |
| **b) Admission Clinical Assessment** |  |  |
| **How long since last admission** |  |  |
| never | 4161 (67.25) | ref |
| <1 month | 693 (11.20) | 1.98 (1.50–2.62) |
| **1 month – 1 year** | 1055 (17.05) | 1.28 (0.96–1.72) |
| **>1 year** | 278 (4.49) | 0.43 (0.17–1.08) |
| **Care sought for current illness prior to admission** | 4103 (66.32) | 1.76 (1.37–2.27) |
| **Self-reported poor prior health** |  |  |
| **Good health prior to this illness** | 5366 (86.73) | ref |
| **<1 week** | 168 (2.72) | 1.42 (0.79–2.54) |
| **1 week – 1 month** | 357 (5.77) | 2.56 (1.84–3.57) |
| **1 month – 1 year** | 296 (4.78) | 3.88 (2.71–5.31) |
| **Referral** | 1883 (30.44) | 2.09 (1.68–2.59) |
| **Prior antibiotic use** | 2732 (44.68) | 1.35 (1.08–1.67) |
| **Prior antimalaria use** | 1425 (23.27) | 1.27 (0.97–1.66) |
| **SpO<sub>2</sub>** | 94.45 (6.45) | 0.96 (0.95–0.97) |
| **SpO<sub>2</sub> <90%** | 944 (15.26) | 1.75 (1.36–2.27) |
| **SpO<sub>2</sub> 90%-95%** | 1448 (23.40) | 0.97 (0.74–1.27) |
| **SpO<sub>2</sub> >95%** | 3795 (61.34) | ref |
| **Heart Rate** | 144.61 (24.82) | 1.00 (0.99–1.00) |
| **Respiratory Rate** | 50.74 (16.71) | 1.01 (1.00–1.01) |
| **Systolic BP** | 92.21 (15.32) | 0.99 (0.99–1.00) |
| **Diastolic BP** | 51.39 (12.59) | 0.99 (0.98–1.00) |
| **Temp (°C)** |  |  |
| **<36.5** | 749 (12.53) | 1.04 (0.75–1.43) |
| **36.5-37.5** | 2965 (49.59) | ref |
| **37.6-39** | 1752 (29.30) | 0.91 (0.71–1.17) |
| **>39** | 513 (8.58) | 0.40 (0.23–0.70) |
| **Respiratory distress** | 1098 (17.75) | 1.46 (1.14–1.87) |
| **Capillary refill ≥3 seconds** | 739 (11.95) | 1.57 (1.19–2.07) |
| **Abnormal Blantyre Coma Scale** | 515 (8.32) | 1.94 (1.44–2.62) |
| **Symptoms during the course of the illness <sup>a</sup>** |  |  |
| **Rash** | 648 (10.47) | 0.72 (0.50–1.06) |
| **Cough <14 days** | 3683 (59.53) | 1.05 (0.85–1.31) |
| Cough >14 days | 462 (7.47) | 1.30 (0.91–1.87) |
| Diarrhoea <14 days | 1877 (30.34) | 0.88 (0.69–1.12) |
| Diarrhoea >14 days | 156 (2.52) | 1.44 (0.81–2.57) |
| Fever <7 days | 5053 (81.67) | 0.79 (0.61–1.03) |
| Fever >7 days | 378 (6.11) | 1.24 (0.83–1.86) |
| Vomiting everything | 1052 (17) | 1.17 (0.87–1.55) |
| Abnormally sleepy | 1247 (20.16) | 1.19 (0.91–1.56) |
| Swelling of both feet | 303 (4.90) | 2.19 (1.51–3.18) |
| Changes in urine color | 707 (11.43) | 1.66 (1.22–2.27) |
| Making less usual than usual | 385 (6.22) | 1.65 (1.17–2.33) |
| Blood in stool | 110 (1.78) | 2.17 (1.24–3.80) |
| Seizure | 876 (14.16) | 1.12 (0.84–1.51) |
| Coma | 42 (0.68) | 2.25 (0.92–5.48) |
| Malaria test positive | 1330 (21.51) | 0.77 (0.57–1.03) |
| HIV RDT test positive | 179 (2.90) | 1.97 (1.24–3.15) |
| Haemoglobin status | 11.87 (3.23) | 0.91 (0.88–0.94) |
| Not anaemic ($\geq 11$g/dL) | 4032 (65.17) | ref |
| Mild anaemia (7-10 g/dL) | 1644 (26.57) | 1.31 (1.03–1.69) |
| Severe anaemia (<7 g/dL) | 511 (8.26) | 2.87 (2.10–3.91) |
| Lactate (mmol/L) | 2.63 (1.91) | 1.08 (1.04–1.14) |
| Glucose (mmol/L) |  |  |
| <2.5 | 154 (2.49) | 1.72 (1.02–2.89) |
| 2.5-8.3 | 5337 (86.26) | ref |
| >8.3 | 696 (11.25) | 1.10 (0.79–1.53) |
| **c) Maternal and social characteristics** |  |  |
| Time to reach hospital |  |  |
| <30 min | 1328 (21.46) | ref |
| 30 min-1h | 2180 (35.24) | 1.65 (1.12–2.42) |
| 1-2h | 1601 (25.88) | 2.92 (2.01–4.24) |
| 2-3h | 737 (11.91) | 2.87 (1.88–4.39) |
| > 3h | 341 (5·51) | 3·81 (2·35–6·18) |
| Distance to facility (km) | 21·40 (21·87) | 1·01 (1·01–1·01) |
| Maternal age (years) | 26·94 (5·81) | 1·01 (0·99–1·02) |
| Household size | 5·02 (2·11) | 1·02 (0·97–1·07) |
| Maternal education | 2·51 (0·85) | 0·69 (0·61–0·79) |
| No school/≤P3 | 632 (10·31) | ref |
| P4-P7 | 2566 (41·88) | 0·72 (0·53–0·98) |
| S1-S6 | 2129 (34·75) | 0·50 (0·36–0·70) |
| Post-Secondary | 800 (13·06) | 0·33 (0·20–0·54) |
| Maternal HIV status |  |  |
| Negative | 5371 (86·81) | ref |
| Positive | 495 (8·00) | 1·53 (1·08–2·17) |
| Unknown | 321 (5·19) | 1·65 (1·09–2·50) |
| Bed net use |  |  |
| Never | 598 (9·67) | 1·19 (0·85–1·68) |
| Sometimes | 473 (7·65) | 1·16 (0·79–1·69) |
| Always | 5112 (82·69) | ref |
| Boil/disinfect/filter water | 4488 (72·54) | 0·75 (0·58–0·97) |
| **d) Discharge characteristics** |  |  |
| Length of stay (days) | 5·41 (7·26) | 1·01 (1·01–1·02) |
| Discharge status |  |  |
| Routine discharge | 5206 (84·17) | ref |
| Referred to higher level of care | 193 (3·12) | 8·81 (6·43–12·06) |
| Unplanned discharge | 788 (12·74) | 3·23 (2·52–4·15) |
| Feeding at discharge |  |  |
| Feeding well | 5707 (92·24) | ref |
| Feeding poorly | 480 (7·76) | 4·43 (3·47–5·65) |
| Discharge Diagnosis |  |  |
| Malaria | 1324 (21·40) | 0·72 (0·53–0·98) |
| Pneumonia | 1926 (31·13) | 1·31 (1·05–1·63) |
| **Bronchiolitis** | 311 (5·03) | 0·70 (0·39–1·25) |
| **Upper respiratory tract infection** | 501 (8·10) | 0·48 (0·27–0·84) |
| **Reactive airway disease/asthma** | 36 (0·58) | 0·57 (0·08–4·07) |
| **Gastroenteritis/diarrhoea** | 964 (15·58) | 0·62 (0·43–0·89) |
| **HIV/AIDS related disease** | 56 (0·91) | 3·06 (1·56–5·97) |
| **Meningitis/encephalitis** | 214 (3·46) | 2·34 (1·56–3·51) |
| **Malnutrition** | 423 (6·84) | 3·21 (2·42–4·26) |
| **Tuberculosis** | 84 (1·36) | 4·37 (2·70–7·07) |
| **Skin/soft tissue infection** | 190 (3·07) | 0·88 (0·46–1·65) |
| **Measles** | 425 (6·87) | 0·39 (0·21–0·73) |
| **Sepsis** | 1762 (28·48) | 0·89 (0·70–1·13) |
| **Genetic/congenital disease** | 115 (1·86) | 5·58 (3·84–8·10) |
| **Sickle cell anaemia** | 62 (1·00) | 0·94 (0·30–2·95) |
| **Febrile seizure** | 25 (0·40) | - |
| **Other: infection** | 133 (2·15) | 1·55 (0·85–2·85) |
| **Other: non-infection** | 236 (3·82) | 4·66 (3·44–6·32) |

All variables adjusted for age, sex and site of enrolment.
SD = standard deviation; aHR = adjusted hazard ratio; CI = confidence interval; MUAC = middle upper arm circumference; BMI = body mass index; SpO<sub>2</sub> = oxygen saturation; BP = blood pressure; Temp = temperature; HIV = human immunodeficiency virus; RDT = rapid diagnostic test; AIDS = acquired immune deficiency syndrome
Note: <sup>a</sup> reference category =no

Regarding factors related to discharge, children with a higher risk of post-discharge mortality included those referred to another hospital for further care (aHR 9·08; 95%CI 6·68–12·34), those with an unplanned discharge (aHR 3·36; 95%CI 2·64–4·28), and those with poor feeding at discharge (aHR 4·53; 95%CI 3·57–5·73). Discharge diagnoses associated with higher post-discharge mortality risk included pneumonia, HIV-related illnesses, tuberculosis, malnutrition, central nervous system infections, and genetic/congenital diseases, while diagnoses associated with lower mortality risk included upper respiratory tract infections, malaria, acute gastroenteritis, and measles (**Table 1d**).

Among variables only collected in the 0-6-month cohort, all measured indicators of acuity were associated with an increased risk of post-discharge mortality, including abnormal tone, pallor, and poor sucking when breastfeeding (**Table S4**).

### Impact of age on post-discharge mortality

Among the four pre-defined age strata, younger age generally conferred a higher risk for post-discharge mortality, although those between 2-6 months of age experienced the highest overall post-discharge mortality rate (**Figure 2**).

**Figure 2.**
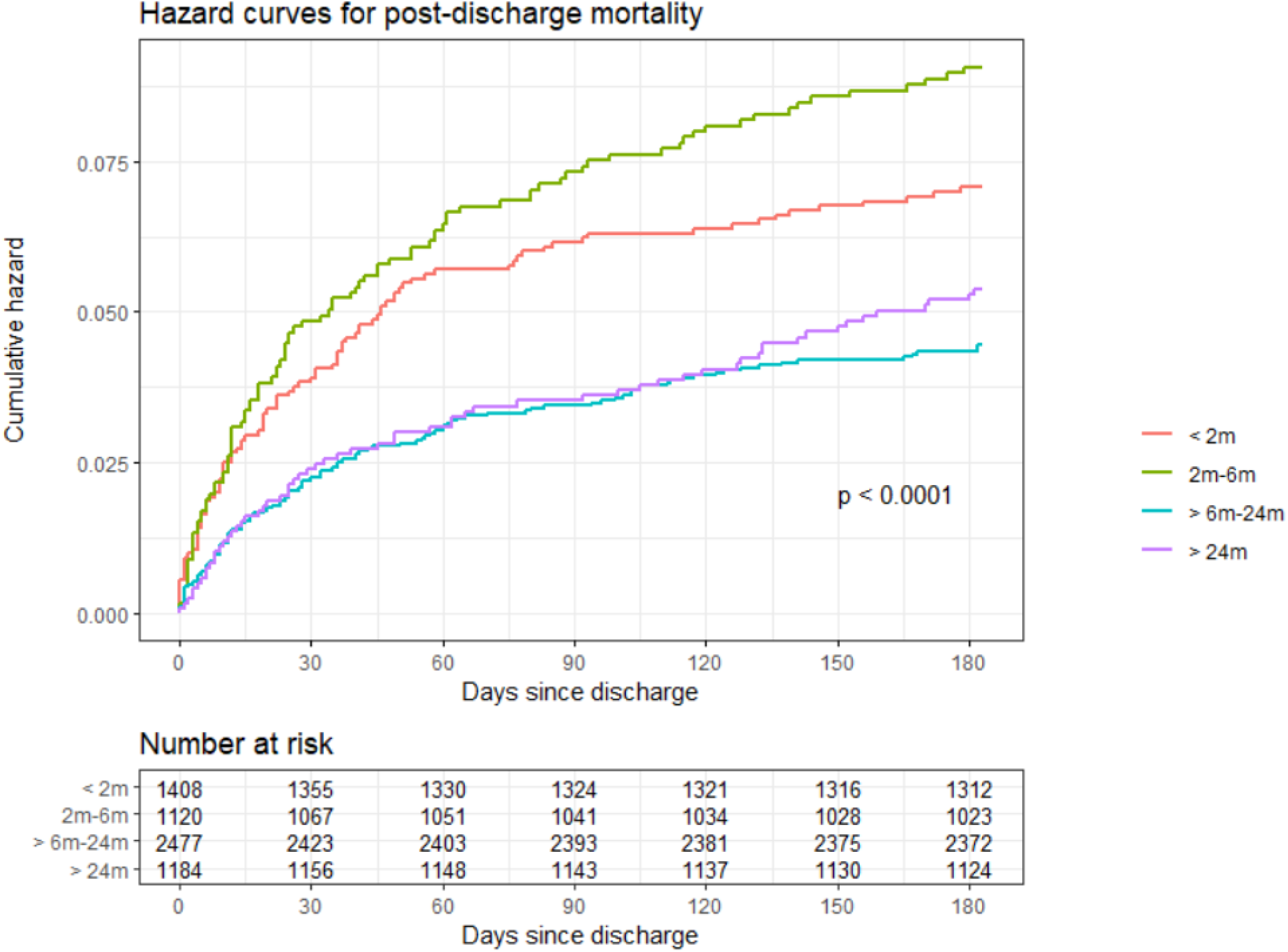
Hazard curves for post-discharge mortality by age category. P-value estimated from log-rank test.

Several clinical parameters measured during hospitalization interacted with age for post-discharge mortality (**Figure 3, Supplementary Figures S1-S6**). As age increased, children presenting with axillary temperatures <36·5^⍰^C experienced a higher probability of post-discharge death. However, other vital signs showed no interactive effect of age, including heart rate, respiratory rate, SpO_2_, and systolic and diastolic blood pressure. Children with severe anaemia appeared to have a consistent risk of mortality across all age strata, while probability of death declined with increasing age among those with moderate or no anaemia. The impact of severely reduced weight-for-age and body mass index (BMI) z-scores appeared to diminish with increasing age, though other anthropometry did not show clear interactive effects. Poor feeding status at discharge and unplanned discharges also disproportionally impacted younger children compared to older children.

**Figure 3.**
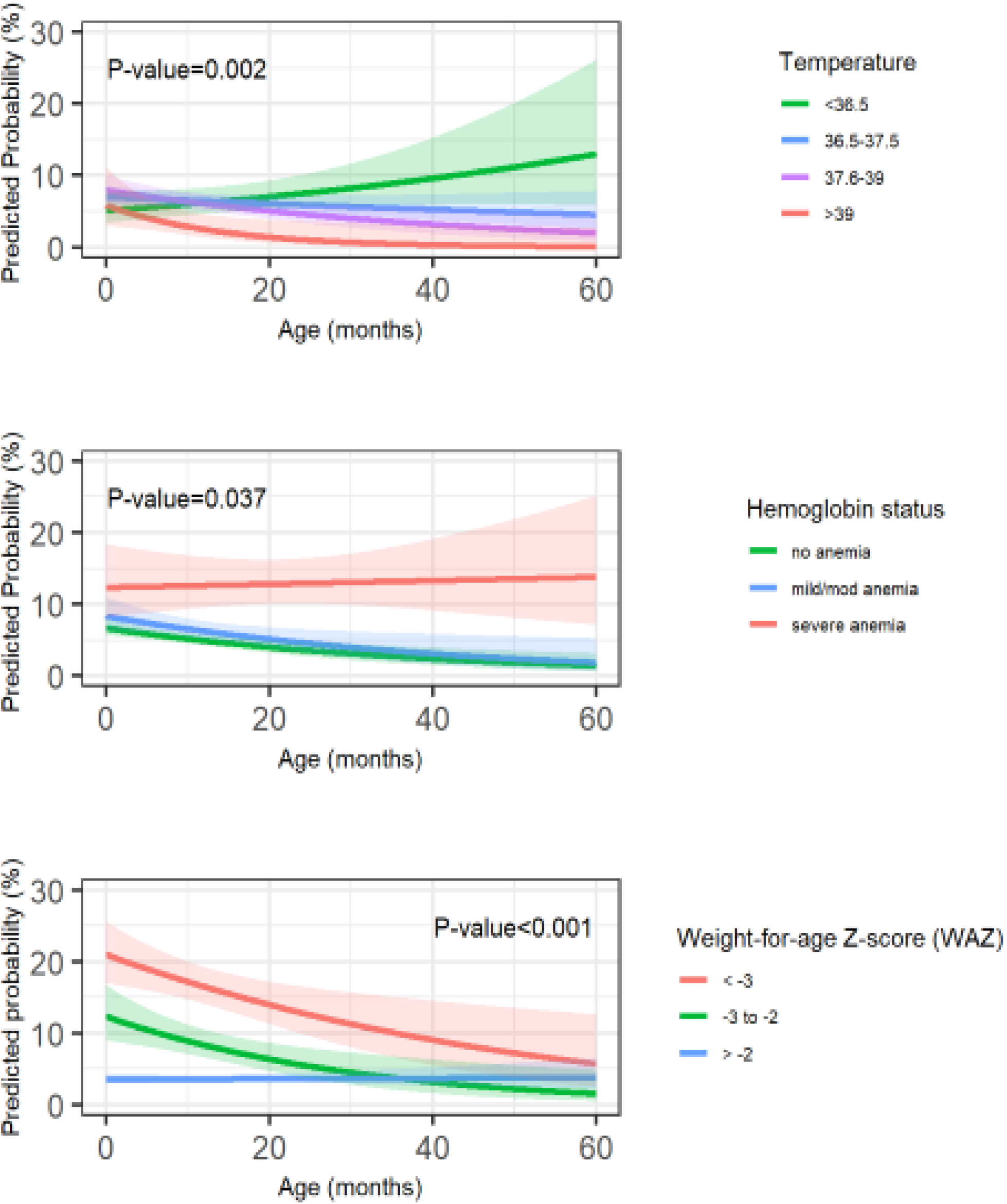
Interactions of age with temperature at admission (top panel), with haemoglobin status at admission (middle panel), and with weight-for-age Z-score (WAZ) at admission (bottom panel). P-value estimated from likelihood ratio test comparing models with and without interaction.

### Timing of post-discharge mortality

The median (IQR) time to post-discharge mortality was 32 (10–92) days from the time of discharge. Using time-period specific hazard ratios corresponding to the tertiles of post-discharge deaths, we observed significant variation among risk factors (**Figure 4, Supplementary Tables S5, S6**). Compared to the hazard ratio of the first period, anthropometry showed persistent risk over the entire period, with peak hazard ratios during the middle tertile. Among laboratory variables, severe anaemia showed clear increasing risk over time. Children with severe anaemia died on average 24 (95%CI 7–40) days later than those without anaemia. The impact of discharge factors including feeding status, referral for further care, and unplanned discharged all diminished significantly over the three post-discharge periods, with as high as a 6-fold reduction in hazard between the first and third periods. Among children who died, those with unplanned discharges died an average of 20 (95%CI 9–32) days earlier than those who were routinely discharged. Social vulnerability, as defined by low maternal education and further distance between home and admitting facility (both in terms of travel time and distance), also demonstrated increasing hazard ratios over the post-discharge time periods.

**Figure 4.**
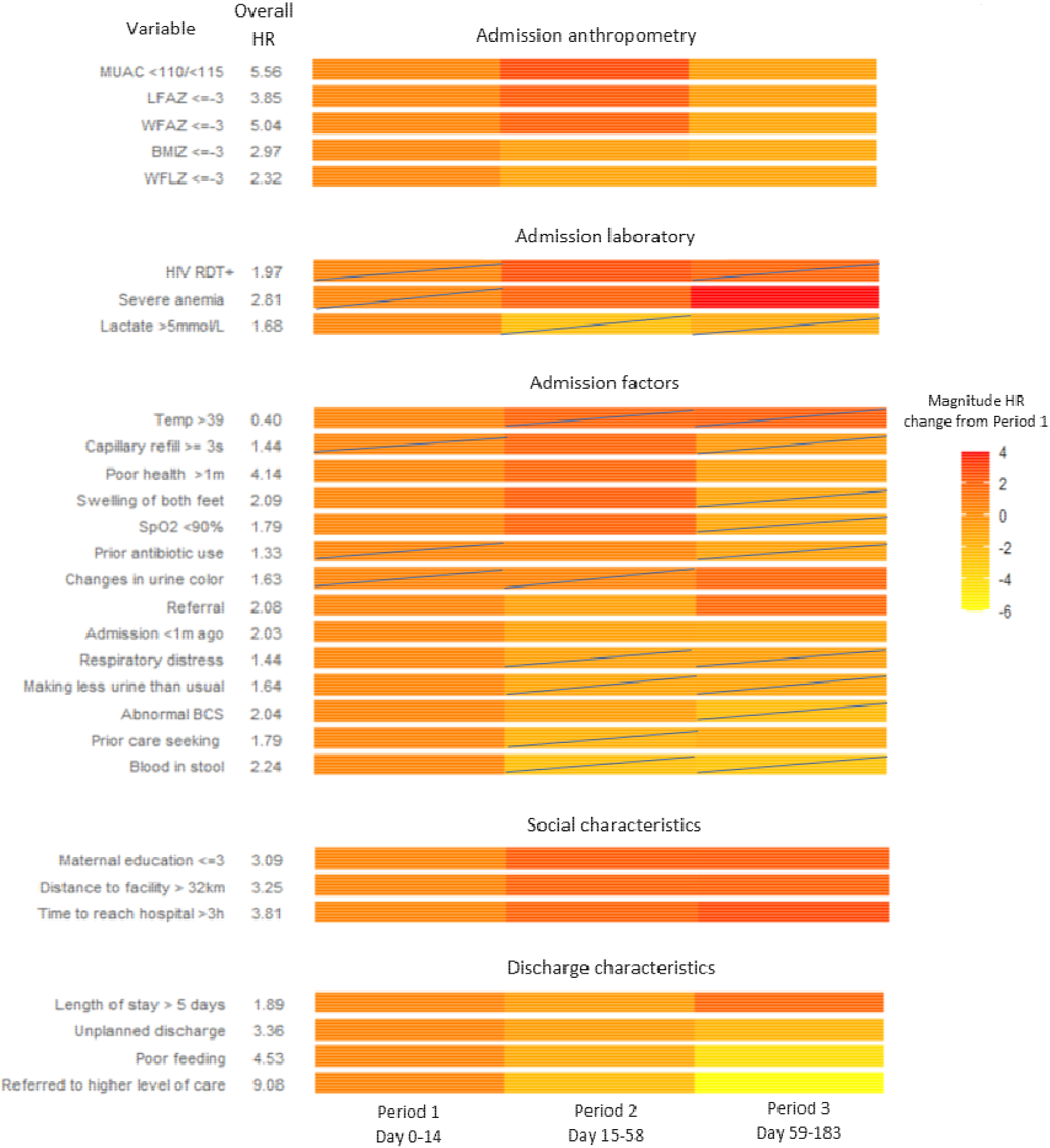
The change in hazard ratio (HR) over time. The 6-month post-discharge period was divided into tertiles according to when deaths occurred. Period specific HRs were calculated. The second and third period colours reflect the magnitude change from the first period according to the legend. Boxes containing diagonal lines represent HRs that are not statistically significant. Actual HR estimates are reported in the supplementary data. *(MUAC = middle upper arm circumference; LFAZ = length for age Z-score; WFAZ = weight for age Z-score; BMIZ = body mass index Z-score; WFLZ = weight for length Z-score; HIV = human immunodeficiency virus; RDT = rapid diagnostic test; Temp = Temperature (oC); SpO2 = oxygen saturation; BCS = Blantyre Coma Scale)*.

### Location of post-discharge mortality

Post-discharge deaths most often occurred at home (n=162, 45%), or in hospital during a readmission (n=132, 37%), though nearly a fifth occurred in-transit while seeking care (n=66, 18%). Neither travel time nor distance from home to the admitting hospital were associated with at-home or in-transit deaths. A discharge diagnosis of malnutrition was associated with at-home deaths, with a relative risk ratio of 1·93 (95%CI 1·03–3·63) compared to in-hospital deaths (**Supplementary Table S7**). The degree of malnutrition was only moderately associated with location of death, with lower age-adjusted mid-upper arm circumference, weight-for-length, and BMI-z scores associated with at-home (but not in-transit) deaths compared to in-hospital deaths.

## Discussion

In this large prospective cohort study of children under-5 years admitted with suspected sepsis, we found that post-discharge mortality was frequent, generally occurred in the first several weeks after discharge, was more common among younger children, and more often occurred in the community than during a readmission. Risk factors for post-discharge mortality, including malnutrition, were largely in line with those reported in previous studies.^4,5,14^ Considering its tremendous impact, strategies to improve care during the hospital-to-home transition must emerge as urgent priorities in research, health policy, and practice.

In these cohorts, nearly half of all post-discharge deaths occurred during the first month of the post-discharge period. The impact of certain risk factors on when children died was notable, especially the increasing risk over time among children with severe anaemia and the decreasing risk over time among children with poor feeding status at discharge and those with unplanned discharges. A child-centred and resource efficient approach to care must therefore ensure that interventions, and their timing, are linked to the at-risk periods. Early mortality could be partially addressed using a discharge readiness checklist, which has demonstrated impact in other settings.^15^ A recent meta-analysis of post-discharge mortality in malaria-endemic areas found that malarial anaemia conferred a lower post-discharge mortality risk compared to non-malarial anaemia,^6^ which was also reflected in our study. A recent study found that post-discharge management of malarial anaemia with malaria chemoprophylaxis following discharge reduced post-discharge readmission and mortality.^16^ However, neither antibacterial chemoprophylaxis (co-trimoxazole) nor iron/folate supplementation appear to improve post-discharge outcomes among children admitted with severe anaemia.^17^

Though younger children tended to have higher rates of mortality after discharge, the highest risk group were those aged 2-6 months. This may be because newborns admitted directly following birth were excluded. The impact of age on certain clinical risk factors was notable, especially with anaemia, temperature, and anthropometry. We found that the impact of anaemia on post-discharge mortality was primarily driven by older children. The contribution of anaemia to post-discharge mortality in older children was recently described in a Tanzanian study,^18^ though it did not include young children and was not large enough to examine age as an interactive factor. The reasons for the interaction we found are unclear and require further investigation. It may be that protective features of foetal haemoglobin and fewer comorbidities among younger children (including chronic malnutrition) play an important role.

Nearly fifty percent of children enrolled in this study had axillary temperatures between 36·5 and 37·5 °C. Higher temperature was associated with lower post-discharge mortality: those with temperatures >39 °C had a hazard ratio of 0·40 (95%CI 0·23–0·70). Furthermore, as children aged, those who died tended to have lower temperatures compared to survivors. These observations are based on predictive axillary surface temperatures and it is unclear whether true core temperatures would differ in their association with post-discharge mortality and interaction with age. Together, these findings point to the need for a more granular understanding of how temperature relates to suspected sepsis.

While the effect of temperature and haemoglobin concentration on mortality increased with age, the impact of weight-for-age and BMI-for-age z-scores appeared to decrease with age. While those who were severely underweight remained at higher risk of death following discharge, the effects of moderate reductions in weight-for-age z-scores (-2 to -3) on mortality largely disappeared. A similar observation was noted in a post-discharge analysis from a surveillance program in Kenya, examining risk factors for children 5-12 years of age, with only severely underweight children experiencing higher risk of death.^19^

Of significant concern are the high proportion of post-discharge deaths (>60%) that occurred in the community. Similar proportions of community deaths have also been observed in community-based death audits, although most caregivers reported having consulted a healthcare provider at some point during the fatal illness.^20^ These observations suggest that health-seeking fatigue over time may be a key factor in community deaths. A nested Kenyan sub-study within the CHAIN network highlighted the often long and complex treatment pathways that acutely ill children navigate: intersecting vulnerabilities at individual, household and facility levels frequently delay and prevent timely and quality care, often resulting in poor outcomes.^21^ A prior analysis of post-discharge deaths in Uganda found that among those who died at home, 90% considered seeking care but could not do so; this was due to a variety of constraints, including financial limitations, lack of transportation, and sudden illness onset.^22^ While it is difficult to disentangle the causal complexity of post-discharge deaths, reducing barriers to accessing existing care in a timely fashion is clearly important. Thus, improving communication and transportation linkages between facilities, and providing scheduled follow-up within communities, especially among vulnerable children, are important strategies that may improve post-discharge outcomes.^23^

## Limitations

This study is subject to some important limitations. Firstly, it was largely exploratory in nature, and included a significant number of analytic comparisons. Additional studies are required to confirm many of these results, and where relevant, should include adjustments for possible confounding variables. In particular, our inclusion criteria, chosen to be generalizable, required only that children had a suspected infection, according to the admitting medical team, and were admitted to hospital; as indicated, these two criteria alone suggest that approximately 90% of these children likely had sepsis according to the IPSCC definition.^10^ Secondly, the study was conducted at sites that were largely rural or semi-rural. Large urban settings are likely to have unique contextual issues compared to rural areas, especially availability of care. However, this may also be a significant strength, as much of the prior literature has focused on urban settings. Finally, this study was conducted in a single country. While the results largely reflect studies conducted in other countries, its geographic limitation may impact its external validity.

## Conclusion

Paediatric post-discharge mortality in the context of suspected sepsis is a common occurrence over the first 6 months, necessitating a robust response among clinicians, researchers, and health policy leaders. Additional resources to improve the hospital-to-home transition are urgently required.

## Data Availability

All data produced in the present study are available upon reasonable request to the authors

## Data sharing

Study materials (including de-identified data, protocol, data collection tools and analysis code) are available upon reasonable request to the corresponding author or through the Pediatric Sepsis CoLab. The University of British Columbia Dataverse Collection: Pediatric Sepsis CoLab. Smart Discharges Dataverse. Borealis. 2022. https://borealisdata.ca/dataverse/smart_discharge

## Author contributions

Contributed to funding applications: MOW, JNB, EK, AT, CB, JS, CO, JMA, NK, JS, CPL, PML, PPM, SN, NKM, JN, JK. Designed the study: MOW, EK, AT, CB, JS, CO, JMA, NK, JS, CPL, PML, PPM, NKM, JK, DD. Coordinated study implementation: MOW, EK, CK, MT, DM, JK. Supervised data collection: SB, AT, SOS, EB, ES, CB, DD, CK, MT, DM. Managed the data: MOW, DD, CK, MT, DM, CZ, MK. Analysed the data: MOW, JNB, CZ, MK, CK, VN. Interpreted the data: MOW, JNB, EK, AT, JMA, NK, JS, CPL, PML, PPM, SN, NKM, NW, VN, JN, JK. Drafted the manuscript: MOW, JNB, NW. Critically reviewed the manuscript: MOW, JNB, EK, SB, AT, SOS, EB, ES, CB, JN, CO, JMA, NK, JS, CPL, PML, DD, PPM, SN, CK, MT, DM, CZ, MK, NW, VN, NKM, JK. All authors and had full access to all the data in the study and accept responsibility to submit for publication.

